# Real-world activity of trastuzumab deruxtecan in heavily pretreated HER2-expressing ovarian cancer: focusing on HER2-low responses and *CCNE1* amplification

**DOI:** 10.64898/2026.06.27.26356757

**Authors:** Gaspar Voelker, Fabian Guelhan, Jannik Luebberstedt, Elisa Schmoeckel, Kai J. Borm, Nicole Pfarr, Maximilian Tschochohei, Leen Houri, Stephanie Fendahl, Elina Arlanch, Marie Koechert, Jonatan Frank, Natyra Tahiri, Alexander Hapfelmeier, Katharina Ilm, Peter Schueffler, Julia Janssen, Martin Boeker, Marion Kiechle, Ulrich A. Schatz, Carolin Mogler, Keno Bressem, Lisa C. Adams, Jacqueline Lammert

## Abstract

**Background:** Trastuzumab deruxtecan (T-DXd) is active in HER2-expressing solid tumours, but trials excluded HER2 immunohistochemistry (IHC) 1+ disease, and data in pretreated ovarian cancer are lacking. We evaluated real-world T-DXd activity and genomic correlates in pretreated ovarian cancer, predominantly high-grade serous (HGSOC).

**Methods:** HER2 expression was assessed in an unselected ovarian cancer cohort (N=74). Fifteen patients receiving off-label T-DXd (14 HGSOC, 1 clear cell; IHC 1+ to 3+) had HER2 status centrally confirmed using gastric-type criteria. Activity was assessed by intra-patient growth modulation index (GMI; progression-free survival [PFS] on T-DXd divided by PFS on the prior line; ≥ 1.33 considered meaningful). Patients on treatment at data cut-off were censored. Objective response (RECIST 1.1) was assessed centrally where imaging was available (n=8).

**Results:** Of the 40 HER2-expressing tumours, 15 received T-DXd, limited mainly by reimbursement. Among 14 evaluable patients (median 5 prior lines), 9 reached a GMI ≥ 1.33 (median 1.69); 8 remained on treatment at cut-off, making durability preliminary. Confirmed partial responses occurred across the HER2 spectrum. Benefit was independent of homologous-recombination (HR) status: one HR-proficient, *CCNE1*-wild-type patient achieved prolonged control and was rendered disease-free after radiotherapy to an oligoprogressive lesion. Exploratory analysis showed all four evaluable *CCNE1*-amplified tumours had reduced or non-durable benefit.

**Conclusions:** T-DXd shows preliminary, clinically meaningful activity in HER2 IHC 1+ ovarian cancer independent of HR status. *CCNE1* amplification may attenuate benefit, a candidate biomarker for WEE1-inhibitor combinations. Approval restricted to IHC 3+ disease would exclude most responders in this cohort. Prospective validation is required.

**Key message:** In heavily pretreated, predominantly HER2-low (IHC 1+) high-grade serous ovarian cancer, trastuzumab deruxtecan showed preliminary, clinically meaningful and homologous-recombination-independent activity. In an exploratory analysis (n=4), *CCNE1* amplification was associated with reduced or non-durable benefit, suggesting a candidate resistance mechanism and a rationale for WEE1-inhibitor combination strategies. Most HER2-expressing patients did not receive treatment, predominantly because of off-label reimbursement barriers rather than clinical ineligibility.

## 1 Introduction

High-grade serous ovarian cancer (HGSOC) carries a poor prognosis in the platinum-resistant setting. Mirvetuximab soravtansine was the first biomarker-directed antibody-drug conjugate (ADC) to confer a survival benefit in platinum-resistant, folate receptor-*α* (FR*α*)-high HGSOC [1]. The high FR*α* eligibility threshold is met in about one third of cases [2]. The larger receptor-low population is a distinct unmet need that may include patients with targetable HER2 expression.

Trastuzumab deruxtecan (T-DXd) is an ADC delivering a membrane-permeable topoisomerase-I payload whose bystander effect provides a rationale for activity in HER2-low tumours. DESTINY-PanTumor02 established activity in HER2-positive solid tumours but explicitly excluded HER2 IHC 1+ disease [3]. DESTINY-Ovarian01 prospectively includes IHC 1+ disease, but only in the treatment-naïve, frontline-maintenance setting in combination with bevacizumab [4]. Real-world data in heavily pretreated, recurrent HGSOC are therefore lacking, and the molecular determinants of payload response and resistance remain poorly defined. We report such a cohort to provide a preliminary signal of clinical activity and to explore genomic correlates of response and resistance, reported in accordance with STROBE (cohort) and REMARK (tumour-marker) recommendations.

## 2 Materials and Methods

### Study design, populations and reporting

This is a multicenter retrospective cohort study of evaluable HGSOC patients and one ovarian clear cell carcinoma patient who underwent central HER2 testing through the molecular tumour board (MTB) at TUM University Hospital. Patients received T-DXd at one of five sites (TUM University Hospital [n=8] and four collaborating regional centres [n=7]) between 22 July 2024 and the data cut-off (i.e. end of the observation period) of 16 June 2026. Receipt of T-DXd and treatment dates for externally treated patients were verified against primary written treatment records. Data were collected within the non-interventional DNPM-Eval framework evaluating MTBs (DRKS00031622). Local ethics approval was granted (No. 2025-362-S-CB); the study followed the Declaration of Helsinki. All patients provided written informed consent to participate in the MTB and for the subsequent use of their clinical and genomic data for research purposes.

Four analysis populations of the 15 treated patients are referred to throughout: all 15 patients (safety and access); 14 patients evaluable for GMI (one began T-DXd shortly before cut-off and is not yet evaluable); 13 patients with comprehensive genomic profiling; and 8 patients with source imaging available for central RECIST 1.1 review.

### HER2 scoring

The Institute of Pathology transitioned its HER2 protocol from breast-cancer to gastric-type criteria during the study period, so the historical screening cohort encompasses both systems; the screening-cohort HER2 prevalence is consequently approximate and likely under-ascertains HER2-low disease. For the 15 patients who received T-DXd, central IHC was uniformly evaluated or retrospectively reassessed by gynaecological pathologists using gastric-type (ISGyP/CAP-gastric) criteria, which account for the incomplete basolateral (“U-shaped”) membranous staining pattern frequently under-scored under strict breast criteria. Reassessment was performed blinded to T-DXd response and clinical outcome. Equivocal cases were reflexed to silver in situ hybridization (HER2 Dual ISH DNA Probe, Ventana).

### Genomic profiling

Comprehensive genomic profiling used the TruSight Oncology 500 (TSO500) DNA panel and the TruSight Tumor 170 RNA panel (Illumina NextSeq550DX or NovaSeq6000). Homologous-recombination deficiency (HRD) was determined from the TSO500 Genomic Instability Score (GIS), standardized to the Myriad myChoice HRD score (positivity threshold GIS ≥ 42) [5]. Variants were classified per ACMG/ESCAT. *CCNE1* amplification was defined as ≥ 5 copies [6].

### Endpoints

Activity was assessed by the intra-patient GMI (PFS on T-DXd ÷ PFS on the preceding progression-defined line; ≥ 1.33 considered meaningful; Von Hoff criterion). The denominator was derived from the most recent treatment line ending in documented progression, treating dose adjustments as continuations and skipping administratively terminated lines in favour of the preceding progression-defined line. Progression was determined by the treating site. For externally treated patients, this was the assessment of the local multidisciplinary tumour board based on routine imaging and was taken as a valid progression endpoint for both the index and reference lines. Patients still on treatment were censored, so their GMI represents a lower bound. Toxicity was graded by CTCAE v5.0.

## 3 Results

### HER2 distribution and access to treatment

In the unselected ovarian cancer cohort (N=74) evaluated for HER2 expression, 34 tumours (46%) were scored HER2 0 and 40 (54%) were HER2-expressing (IHC 1+ to 3+); because the screening cohort was scored under heterogeneous criteria, these proportions are approximate. Of the 40 HER2-expressing patients, 15 (38%) received off-label T-DXd (Figure 1). Among the 25 untreated patients, documented reasons were ongoing standard therapy or trial enrolment (10), rapid clinical deterioration (9), a denied or pending off-label cost-coverage application (4), failure to initiate the application (1), and a competing second malignancy (1). T-DXd had no approved ovarian or tumour-agnostic EU indication during the study, so every patient, including IHC 3+ cases, was treated off-label. Because a treated-versus-untreated comparison would be confounded, we report the cascade descriptively and assess activity by intra-patient GMI.

**Figure 1:**
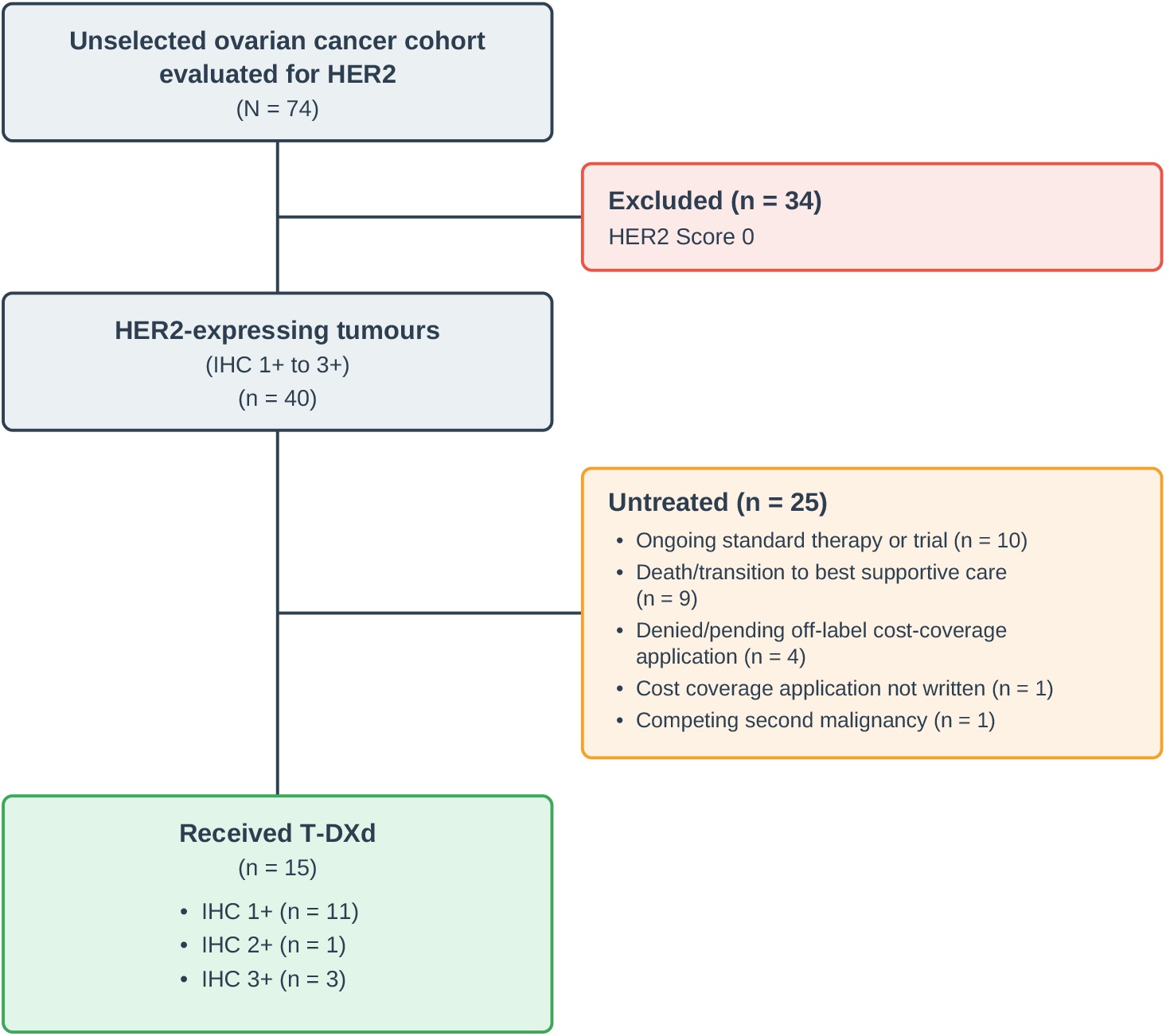
Patient flow diagram showing disposition and access to T-DXd. The diagram shows screening, identification and attrition of patients evaluated for off-label T-DXd. From an unselected ovarian cancer cohort (N=74), patients without HER2 expression (score 0; n=34) were excluded. Of 40 HER2-expressing tumours (IHC 1+ to +), 25 did not receive T-DXd the diagram details the clinical trajectories and administrative barriers involved. The treated cohort (n=15) is stratified by central HER2 IHC score.

### Cohort and activity

The 15 treated patients comprised IHC 1+ (n=11), IHC 2+ (n=1) and IHC 3+ disease (n=3); 14 were high-grade serous and one ovarian clear cell carcinoma (HER2 IHC 1+). Median age at T-DXd initiation was 70 years (range 38–85), after a median of 5 prior systemic lines (range 2–7), with T-DXd given between the 3rd and 8th line; five had received prior mirvetuximab (Table 1).

**Table 1.**
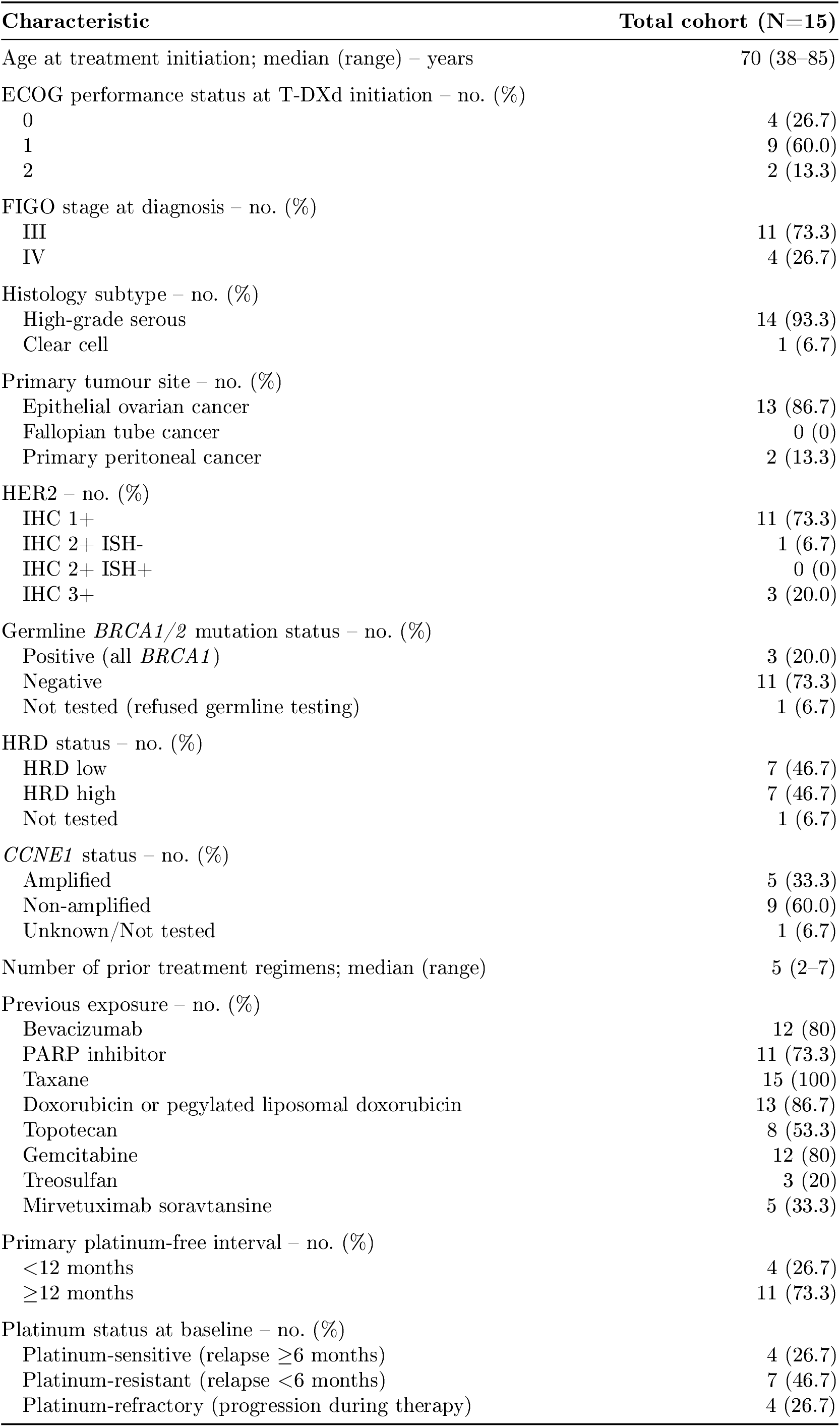
Baseline clinical, demographic, and genomic characteristics of the treated cohort (N=15).

At least 9 of 14 evaluable patients reached a GMI ≥ 1.33 (median 1.69) – a notable signal, since a GMI *>* 1 is progressively harder to achieve (Figure 2; individual treatment trajectories in Supplementary Figure 1). Eight of 15 remained on treatment at cut-off, and six of these had initiated T-DXd within roughly four months of the data cut; many GMI values are therefore lower bounds, and durability must be read as an early sign. Of the five evaluable values below the 1.33 threshold, three were *CCNE1*-amplified tumours without durable benefit (below), one a censored lower bound in a patient still early on treatment, and one a partial responder who progressed after a duration comparable to her preceding line.

**Figure 2:**
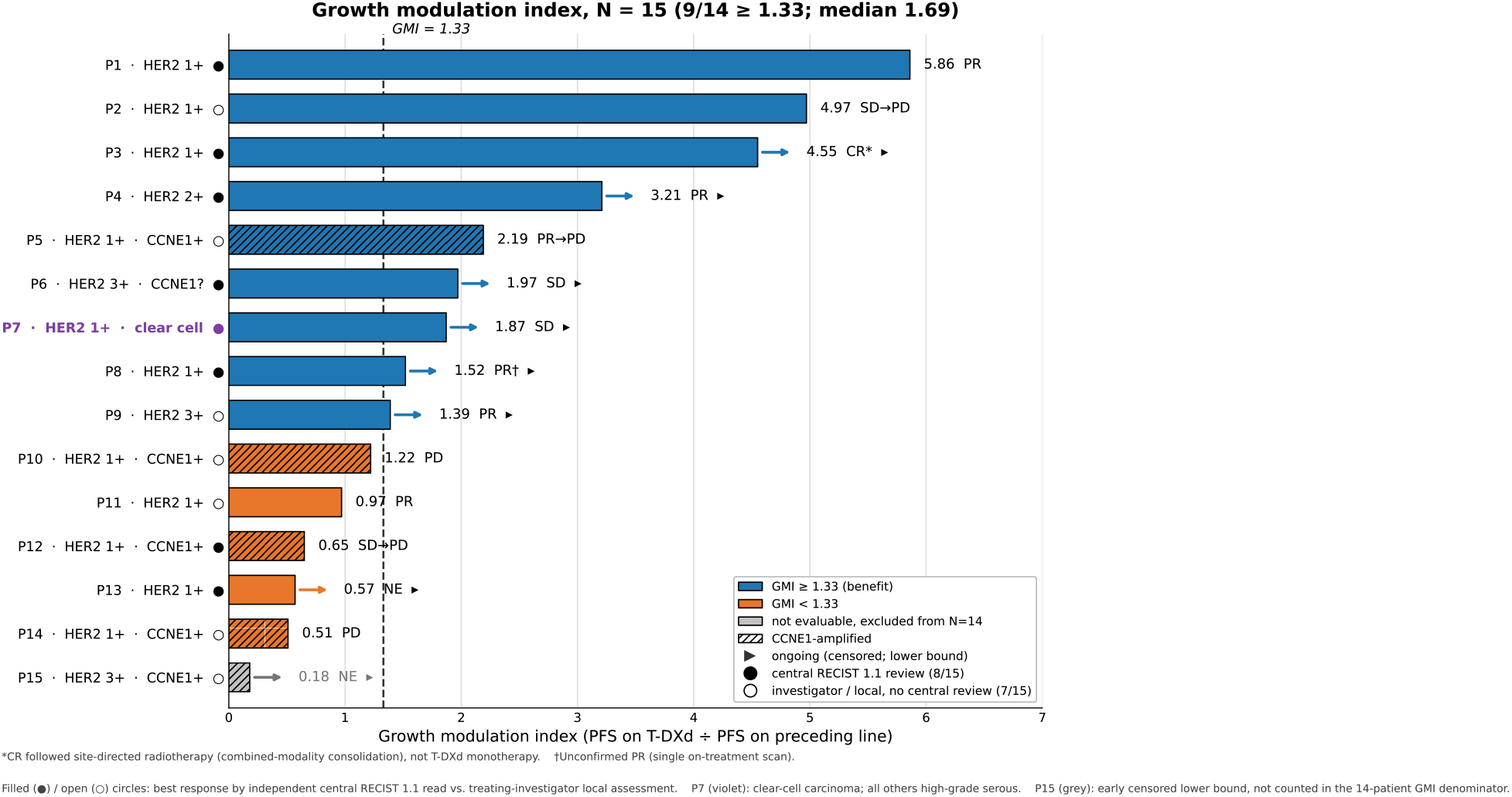
Intra-patient growth modulation index (GMI). Bars show the GMI for each treated patient (PFS on T-DXd divided by PFS on the immediately preceding progression-defined line). A GMI ≥ 1.33 indicates clinically meaningful benefit. Black triangles denote patients on treatment at data cut-off, whose GMI is a censored lower bound; several of these patients had only a few months of exposure, so their values are provisional. Striped bars denote *CCNE1*-amplified tumours; note that one *CCNE1*-amplified patient exceeded the 1.33 threshold despite a non-durable partial response, so the colour threshold should not be read as durable benefit. Best overall responses are annotated adjacent to each bar.

### Objective responses

Central RECIST 1.1 re-read was available for 8 of 15 patients (Supplementary Table 1); among the 7 RECIST-evaluable (one not yet evaluable), 3 achieved a confirmed partial response spanning the HER2 expression spectrum, and a fourth had a radiotherapy-consolidated complete response reported separately as a combined-modality outcome.

Lesion-level tracking revealed mixed responses even in patients with overall stable disease. For example, one patient showed shrinkage of the primary tumour alongside slight progression of a single liver lesion, suggesting that early discontinuation on heterogeneous lesion behaviour may be premature in this population. The most durable benefit occurred in a HER2 IHC 1+ (HRD-negative, *BRCA1/2*-wild-type, *CCNE1*-wild-type). This patient has remained on T-DXd for approximately 23 months and, after an episode of localised progression treated with site-directed radiotherapy, had no evidence of disease on two consecutive PET-CT assessments. Because radiotherapy was delivered to the progressing lesion, this complete response is a combined-modality outcome, described as durable disease control on T-DXd consolidated by ablative radiotherapy.

### Safety

Haematological events predominated (anaemia, thrombocytopenia, and leukopenia/neutropenia Reported gastrointestinal and systemic toxicity was lower than in pivotal trials, likely reflecting retrospective under-ascertainment and mandatory triple-antiemetic prophylaxis. One patient experienced grade 3 metabolic acidosis. No clinically significant fall in left ventricular ejection fraction and no drug-related interstitial lung disease (ILD) were documented; given the small cohort and the approximately 10% drug-related ILD rate in DESTINY-PanTumor02 [3], this absence should not be read as a safety signal.

### Genomic associations (exploratory)

Comprehensive genomic profiling was available for 13 of 15 treated patients. Clinical benefit was independent of homologous-recombination status: all three germline *BRCA1*-mutant patients responded, an *ARID1A*-mutant tumour achieved disease control (SD), and the most durable responder was confirmed HRD-negative and wild-type for *BRCA1/2, ARID1A* and *CCNE1*, consistent with HR-independent payload and bystander cytotoxicity. *CCNE1* amplification was present in five tumours (four IHC 1+, one IHC 3+ [not yet evaluable]). Of the four evaluable IHC 1+ tumours, none derived durable benefit: three showed no meaningful benefit (GMI 0.5–1.2), including one early death after a single dose that reflected high disease burden rather than molecular escape, and the fourth (HRD-low) achieved a RECIST partial response that was not sustained, progressing within weeks of the initial assessment.

## 4 Discussion

In this real-world cohort, T-DXd showed preliminary, clinically meaningful activity in heavily pretreated, HER2-expressing ovarian cancer, even in HER2-low (IHC 1+ or IHC 2+/ISH-) disease, which, as presented, constitutes a substantial proportion of patients. However, our findings highlight a critical biological divergence: while *CCNE1* wild-type tumours can achieve durable, sustained response, tumours harbouring *CCNE1* amplification show rapid loss of benefit from the topoisomerase-I payload. We present this as a hypothesis rather than an established effect; the subgroup is small and one case was confounded by early death due to rapid disease progression, but it points to a specific, potentially targetable escape mechanism in a population that is excluded from pivotal trials and largely without approved access.

The overall benefit we observed occurred irrespective of homologous-recombination (HR) status, a finding that merits emphasis because it separates T-DXd from the DNA-repair-directed logic of PARP inhibition that dominates ovarian-cancer therapeutics [7, 8]. The DXd payload is an intracellularly delivered topoisomerase-I inhibitor; it is highly membrane-permeable, enabling the bystander killing of neighbouring antigen-low cells [9, 10]. Because T-DXd delivers this payload directly into the cell at a high drug-to-antibody ratio, the resulting targeted DNA damage can overwhelm even fully functional HR mechanisms – neither property requires a pre-existing DNA-repair defect to be effective.

In our cohort, germline *BRCA1*-mutant and *BRCA*-wild-type tumours both responded, an *ARID1A*-mutant tumour responded, and the most durable course arose in a confirmed HRD-negative, *BRCA1/2*-wild-type tumour. This pattern is consistent with payload- and bystander-driven cytotoxicity operating across HR backgrounds and implies that HR status – central to selecting PARP-inhibitor candidates – is unlikely to be a useful selection biomarker for T-DXd. The caveat is that the deepest response was consolidated by site-directed radiotherapy and is therefore a combined-modality outcome; it illustrates rather than proves HR-independent durability, and the aggregate of HR-deficient and HR-proficient responders is the stronger basis for the inference.

### Mechanistic rationale (preclinical hypothesis)

The pattern we observed is mechanistically coherent with published preclinical work, which we summarise to frame a testable hypothesis rather than to assert that the mechanism is operative in our patients. The DXd topoisomerase-I payload induces DNA double-strand breaks preferentially during S-phase. *CCNE1*-amplified tumours, driven by cyclin E1 overexpression, undergo premature S-phase entry and carry high baseline replication stress; to resolve payload-induced damage before mitosis they become disproportionately dependent on the G2/M checkpoint [11]. In HER2-low, *CCNE1*-amplified xenografts, T-DXd monotherapy produces only temporary growth inhibition – reminiscent of the transient responses in our *CCNE1*-amplified patients – whereas adding a WEE1 inhibitor (adavosertib in those models) abrogates the G2/M checkpoint and drives near-complete regression [11]. This sensitisation is not seen in HER2-overexpressing (IHC 3+) models, which respond to T-DXd alone [11]. An analogous clinical association – shorter PFS in *CCNE1*-altered HER2-positive gastric cancer (reported median 131 vs 189 days; hazard ratio 1.90, 95% CI 1.02–3.53) [12] – provides external, hypothesis-supporting context; it is not evidence generated in our cohort, and the gastric setting differs from ovarian disease.

We distinguish the preclinical agent (adavosertib) from the clinical agent now in trials (azenosertib); both are WEE1 inhibitors, but the xenograft regression data were generated with adavosertib and should not be read as clinical validation of any specific compound.

Taken together, these strands imply that future evaluation of T-DXd in HER2-low ovarian cancer should account for cyclin E status rather than relying on HER2 expression alone. If the preclinical model holds, cyclin-E-dysregulated tumours – defined by *CCNE1* amplification and/or cyclin E1 protein overexpression – would be the rational population in which to test the addition of a WEE1 inhibitor, whereas cyclin-E-wild-type tumours might derive durable benefit from T-DXd alone. This remains a hypothesis generated by four amplified cases, one of them confounded by early death; we raise it because the underlying biology is specific and testable, and because the population it identifies is currently neglected. The recurrent HER2 IHC 1+ setting is addressed by DESTINY-Ovarian01 only as frontline maintenance. The ongoing phase I of T-DXd plus a enosertib 13 does admit HER2-low disease (IHC 1+/2+), but administers the combination to all HER2-expressing patients irrespective of *CCNE1* status – cyclin E is assessed only as an exploratory biomarker – and confines its efficacy expansion to gastric/gastroesophageal-junction disease. Consequently, no current trial tests a *CCNE1*-stratified strategy in ovarian cancer, nor whether *CCNE1*-wild-type tumours derive durable benefit from T-DXd monotherapy without the added toxicity of WEE1 inhibition. Our cohort is intended to justify, not to substitute for, prospective and appropriately biomarker-stratified evaluation.

### Reconciling the IHC 1+ findings, histology and sequencing

Our IHC 1+ observations must be reconciled with emerging real-world series that have not reported objective responses at IHC 1+. In one cohort of 33 HER2-expressing ovarian and endometrial cancers, T-DXd produced no objective responses among IHC 1+ tumours yet achieved 100% disease control with a median of 7.3 months on treatment [14]. We read this as concordant rather than contradictory: the dominant benefit we observed at IHC 1+ was within-patient disease stabilisation captured by GMI, matching their high disease-control rate and prolonged treatment durations. Our confirmed partial responses are the additional observation, interpreted cautiously given immature follow-up and central re-read in only a subset. Central, uniform gastric-type scoring – which recovers incomplete basolateral staining missed by breast criteria – is one plausible contributor, supported by the central HER2 reassessment in the DESTINY-PanTumor02 gynaecological analysis [15]; whether centrally scored IHC 1+ disease yields reproducible objective responses will require prospective, centrally read validation. Histology appears to constrain benefit: the largest ovarian series reported uniform resistance in clear cell carcinoma, with responses concentrated in high-grade serous and mucinous disease [16], and the only prior exceptional response was HER2 IHC 3+ [17]. While our single *ARID1A*-mutant clear cell case achieved SD despite being HER2 IHC 1+, no definitive conclusion can be drawn from a single patient, and this histology warrants dedicated evaluation in larger, stratified cohorts. ADC sequencing also remains open: durable T-DXd activity after sacituzumab govitecan has been reported [18], and one patient in our cohort, having progressed on T-DXd, is now receiving the reverse sequence – a prospective opportunity to test whether switching the target antigen overcomes resistance to a shared payload class.

### Access and implementation

The number of patients reaching treatment is itself a central message. Of 40 HER2-expressing tumours identified through the DNPM-Eval framework, only 15 (38%) received T-DXd. This implementation gap, driven predominantly by off-label reimbursement barriers rather than clinical ineligibility, is precisely the access-to-implementation shortfall the DNPM evaluation was designed to capture, and it localises a modifiable failure point in the care pathway. Four patients had cost-coverage applications denied or left pending and died before or shortly after a decision. We are deliberately cautious about causal interpretation here: this outcome is consistent with a harmful barrier, but also with confounding by indication and immortal-time bias, since the sickest patients are least able to await a decision. What can be said without that confound is structural: in May 2026 the EMA’s CHMP recommended the first tumour-agnostic HER2-directed indication in the EU [19], but – like the US approval – restricted to HER2 IHC 3+ disease, leaving the IHC 2+ and large IHC 1+ subgroups without an approved option, and leaving the real-world genomic datasets needed to validate resistance mechanisms such as the *CCNE1* signal reported here dependent on ad hoc off-label access.

### Limitations

This is a small, retrospective cohort without a randomised comparator. HER2 scoring was central; treatment delivery, progression assessment and toxicity capture were performed at the treating sites by local multidisciplinary tumour boards, with central RECIST 1.1 re-read available for 8 patients – reflecting routine real-world practice rather than blinded central trial review. Follow-up is immature, several patients had only a few months of T-DXd exposure at cut-off, and a substantial fraction of GMI values and responses are censored or not yet confirmed. The heterogeneous historical HER2 scoring in the untreated screening cohort means the prevalence of HER2-low disease is approximate and probably under-represented. The genomic analysis is strictly exploratory, the *CCNE1*-amplified subgroup is small, and untreated HER2-expressing patients are not a valid outcome comparator owing to deep confounding. Larger, prospective, biomarker-stratified validation is required, including evaluation of *CCNE1* amplification as a modifier of T-DXd benefit, particularly in combination with WEE1 inhibition.

In conclusion, T-DXd showed preliminary, HR-independent activity in heavily pretreated HGSOC enriched for HER2 IHC 1+ disease. *CCNE1* amplification was associated with reduced benefit and provides a strong biological rationale for WEE1-inhibitor combination strategies. Wider adoption of disease-specific HER2 scoring and removal of off-label access barriers are prerequisites for delivering and rigorously evaluating HER2-directed ADC therapy in ovarian cancer.

## Acknowledgements

This work is supported by the Bavarian State Ministry of Health, Care and Prevention through the GO-TWIN project (grant number DGP-2024-01).

## Data availability

All data needed to evaluate the conclusions of this study are present in the article and its Supplementary Information. Individual patient-level clinical and raw sequencing data are not publicly available because they could compromise patient privacy and fall outside the consent and ethics terms governing this retrospective study (DRKS00031622; ethics No. 2025-362-S-CB), but de-identified data are available from the corresponding author on reasonable request for non-commercial academic research.

## Author contributions

Conceptualisation: G.V., J.L.; project administration: J.L.; resources: F.G., J.Lu., E.S., K.J.B., N.P., M.T., L.H., S.F., J.F., K.I., U.A.S., C.M., K.B., L.A. and J.L.; validation: G.V., E.S., N.P., E.A., M.K., N.T., K.I., U.A.S., C.M.; writing – original draft preparation: G.V., J.L.; writing – review & editing: F.G., J.Lu., E.S., K.J.B., N.P., M.T., L.H., S.F., E.A., M.K., J.F., N.T., A.H., K.I., J.J., P.S., M.B., M.Ki., U.A.S., C.M., K.B., L.A., J.L. All authors contributed scientific advice and approved the final version of the manuscript.

## Supplementary information

**Supplementary Figure 1:**
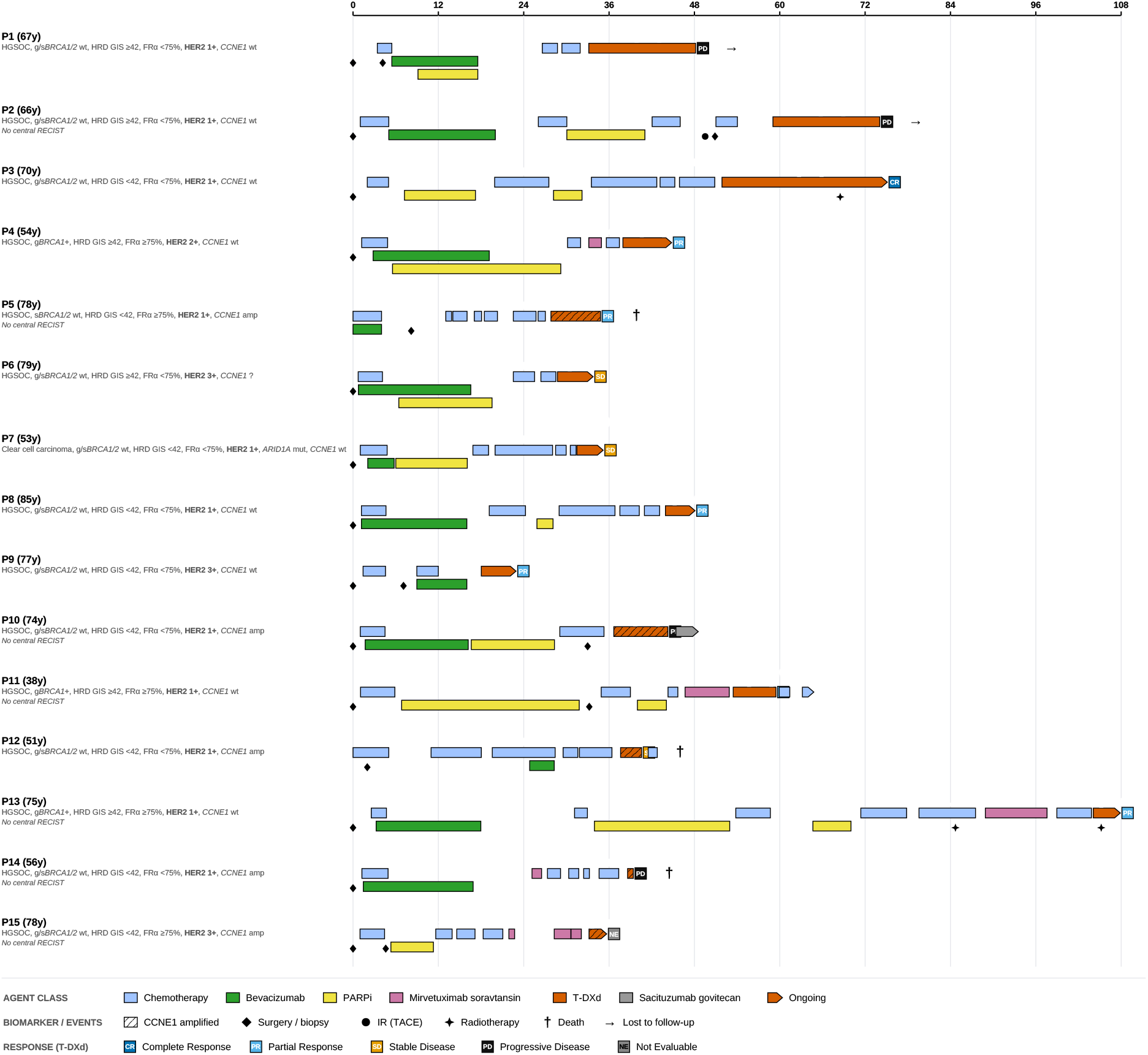
Comprehensive longitudinal treatment trajectories and biomarker profiles for the T-DXd cohort. Swimmer plots detail the individual therapeutic histories for all 15 patients from initial diagnosis through to the data cut-off. The vertical ordering and numbering of patients (P1–P15) correspond directly to the GMI ranking shown in Figure 2. Hatched T-DXd bars denote tumours with *CCNE1* amplification. Key clinical events (surgeries, biopsies, site-directed radiotherapy, death or loss to follow-up) are marked with distinct symbols; best objective responses on T-DXd are indicated, and pointed arrowheads denote treatments ongoing at data cut-off.

**Supplementary Table 1:**
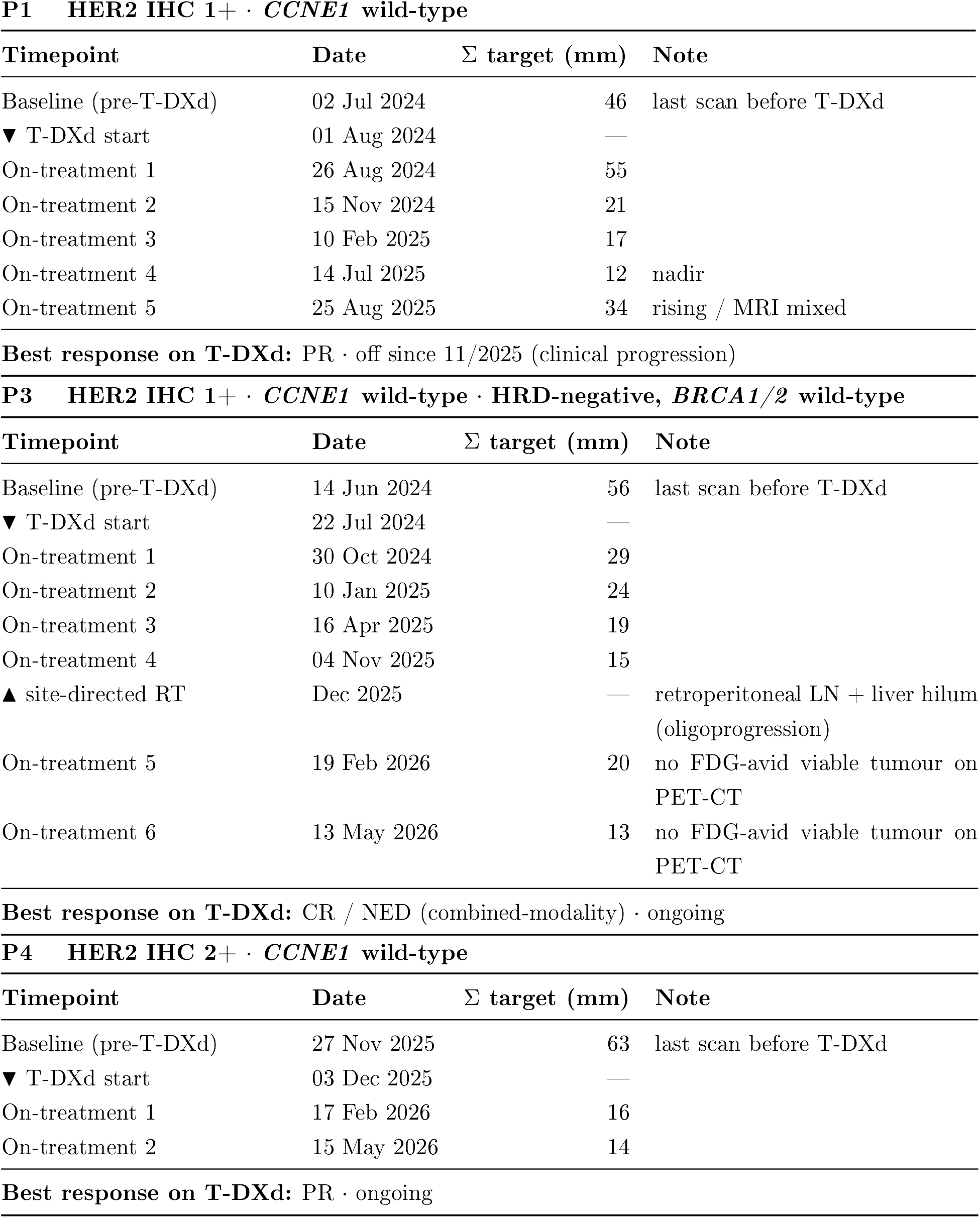
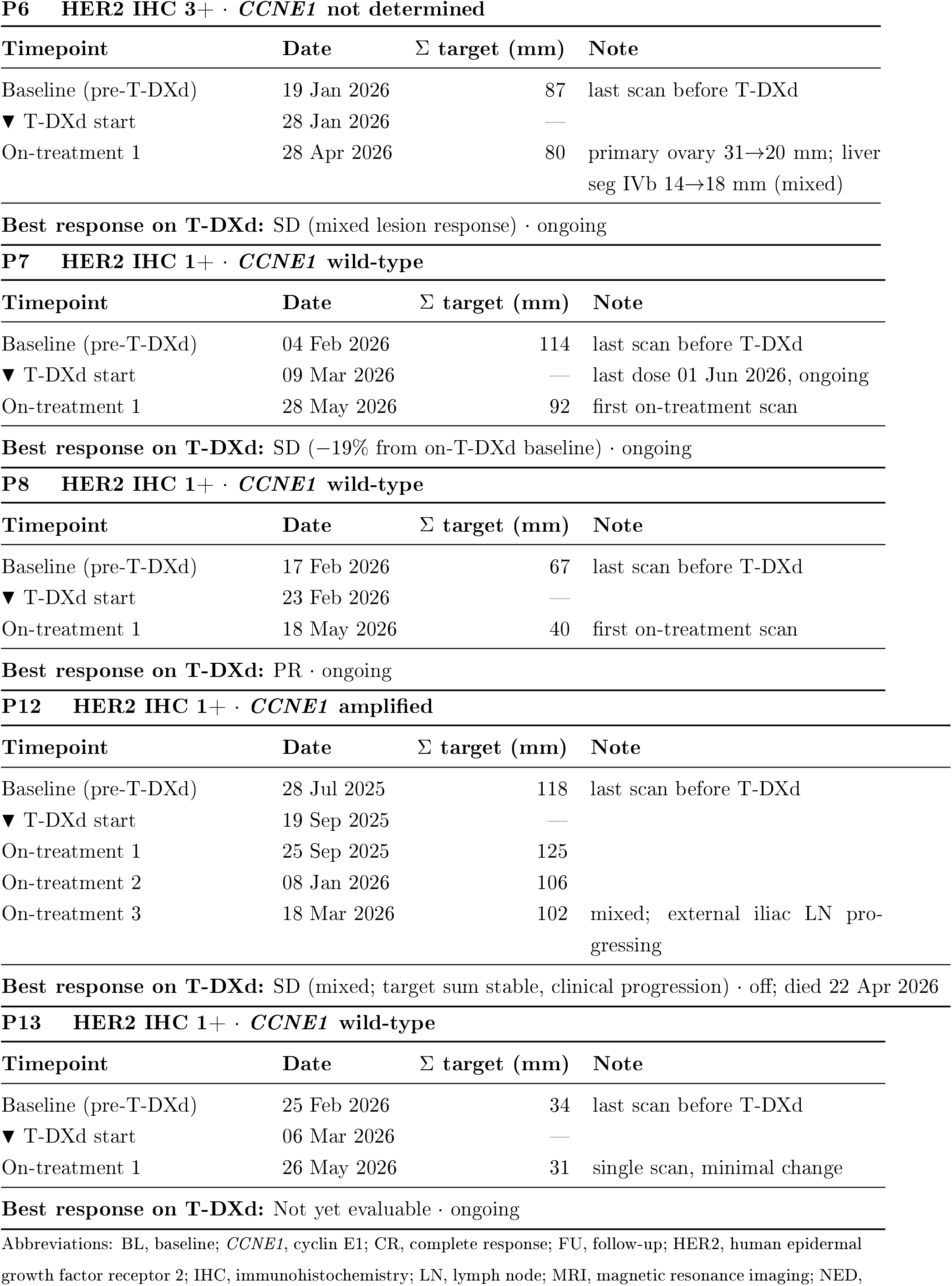
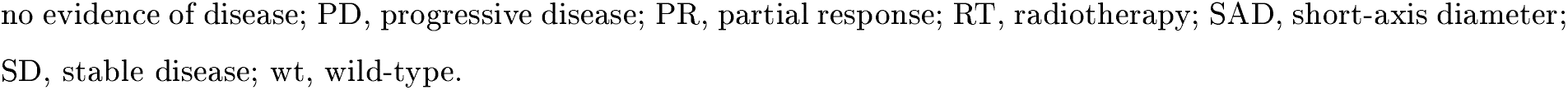
Longitudinal radiographic tumour response. Central imaging evaluation of the cohort (n=8; 7 high-grade serous, 1 clear cell). The “▼ T-DXd start” marker denotes trastuzumab deruxtecan initiation, with baseline defined as the most recent scan prior to this marker; the “▲ site-directed RT” marker denotes intercurrent radiotherapy. Tumour response was assessed per RECIST 1.1. Values represent the sum of target-lesion diameters (mm; short-axis for lymph nodes). Best response on T-DXd is derived exclusively from post-treatment scans; patients with insufficient follow-up are noted as not yet evaluable.

